# Determinants and trends of COVID-19 vaccine hesitancy and vaccine uptake in a national cohort of U.S. adults: A longitudinal study

**DOI:** 10.1101/2021.05.12.21257116

**Authors:** Madhura S. Rane, Shivani Kochhar, Emily Poehlein, William You, McKaylee M. Robertson, Rebecca Zimba, Drew A. Westmoreland, Matthew L. Romo, Sarah G. Kulkarni, Mindy Chang, Amanda Berry, Angela M. Parcesepe, Andrew R. Maroko, Christian Grov, Denis Nash, the CHASING COVID Cohort Study Team

## Abstract

We estimated the trends and correlates of vaccine hesitancy, and its association with subsequent vaccine uptake among 5,458 adults in the United States. Participants belonged to the CHASING COVID Cohort, a national longitudinal study. Trends and correlates of vaccine hesitancy were examined longitudinally in eight interview rounds from October 2020 to July 2021. We also estimated the association between willingness to vaccinate and subsequent vaccine uptake through July 2021. Vaccine delay and refusal decreased from 51% and 8% in October 2020 to 8% and 6% in July 2021, respectively. Compared to Non-Hispanic (NH) White participants, NH Black and Hispanic participants had higher adjusted odds ratios (aOR) for both vaccine delay (aOR: 2.0 [95% CI: 1.5, 2.7] for NH Black and 1.3 [95% CI: 1.0, 1.7] for Hispanic) and vaccine refusal (aOR: 2.5 [95% CI: 1.8, 3.6] for NH Black and 1.4 [95% CI: 1.0, 2.0] for Hispanic) in June 2021. COVID-19 vaccine hesitancy was associated with lower odds of subsequent vaccine uptake (aOR: 0.15, 95% CI: 0.13, 0.18 for vaccine-delayers and aOR: 0.02; 95% CI: 0.01, 0.03 for vaccine-refusers compared to vaccine-willing participants), adjusted for sociodemographic factors and COVID-19 history. Vaccination awareness and distribution efforts should focus on vaccine delayers.

---

As the coronavirus disease-19 (COVID-19) pandemic continues to be a health crisis globally, widespread vaccination is the most effective and sustainable long-term mitigation strategy. Thirteen safe and efficacious vaccines were developed and authorized worldwide within a span of a year since the World Health Organization (WHO) declared the COVID-19 outbreak a pandemic (1). In the United States (U.S.), as of September 2021, the Pfizer-BioNTech BNT162b2 vaccine (2) is fully approved for adults, while the Moderna mRNA-1273 (3) and the Janssen Ad26.COV2.S vaccines (4) are currently authorized for emergency use. The Pfizer-BioNTech and the Moderna vaccines are both 2-dose mRNA vaccines, while the Janssen vaccine is a single dose, non-replicating viral vector vaccine. Among U.S. residents 12 years and older, 62% have received at least one dose and 52.7% have been fully vaccinated as of September 2, 2021; however vaccination rates vary by state and county and demand for coronavirus vaccines has decreased in recent months (5,6). As vaccine eligibility criteria expands, we would expect dramatic reductions in COVID-19 incidence, hospitalizations, and mortality in all age groups as vaccine uptake increases (7).

For the COVID-19 vaccination program to be as impactful as possible, large numbers of people must be vaccinated quickly while also ensuring equity in access and uptake. Low vaccine acceptance and lack of easy access to vaccinations can be barriers to achieving both high and equitable vaccination coverage (8). This could create vaccination cold-spots where periodic disease outbreaks can still occur (9) and vaccine-resistant strains might evolve (10). While anti-vaccine sentiment remains a threat to COVID-19 vaccine uptake in the U.S., other factors, such as political mistrust, lack of assurance about safety and efficacy, and a lack of clear public health messaging may have influenced vaccine hesitancy specifically for coronavirus vaccines (11). The rapid production of COVID-19 vaccines in less than a year may have engendered concerns among the public, considering the average vaccine development timeline spans around ten years (12). According to a Kaiser Family Foundation poll from August 2020, a majority (62%) of respondents believed that socio-political factors and pressures could lead to a rushed approval for the COVID-19 vaccine without assurances of safety and efficacy, and only 42% of the participants were willing to get the COVID-19 vaccine if approved before the U.S presidential elections in November 2020 (13). Based on a systematic review of surveys conducted between April and October 2020, the U.S. recorded lower intention to vaccinate against COVID-19 (ranging from 38-49% across regions) compared to other high-income countries such as Denmark (80%) and the United Kingdom (79%) (14).

Understanding COVID-19 vaccine hesitancy and addressing it promptly is essential for a successful and equitable vaccine roll-out. In this study, we aimed to 1) measure trends in vaccine hesitancy in the U.S. for adults; 2) identify subpopulations that might be less willing to be vaccinated; 3) examine sociodemographic and behavioral factors as well as COVID-related risk perceptions that correlate with vaccine hesitancy; and finally, 4) assess the association between vaccine hesitancy and subsequent vaccine uptake.

## METHODS

### Study design and participants

This study used data from the Communities, Households, and SARS-CoV-2 Epidemiology (CHASING) COVID Cohort study. CHASING COVID is a national prospective cohort study in the U.S. launched on March 28, 2020, to understand the spread and impact of the SARS-CoV-2 pandemic within households and communities. Details of survey methodology are described elsewhere (15). Briefly, study participants were recruited through social media platforms or through referrals using advertisements that were in both English and Spanish. Eligible participants were ≥18 years old U.S. residents with a valid ZIP code and email address. Patient consent for survey participation and serological testing was obtained. As of September 7, 2021, eight full interview rounds (March 2020 [V0], April 2020 [V1], July 2020 [V2], October 2020 [V3], November 2020 [V4], December 2020 [V5], February 2021[V6], June 2021 [V7]) were completed which captured longitudinal information on participant demographics, COVID-related exposures, outcomes, detailed symptoms, non-pharmaceutical intervention (NPI) use, vaccine uptake, and other behavioral factors. Additionally, 3 shorter interviews (April 2021[V6.1], May 2021 [V6.2], July 2021 [V7.1]) were administered to capture COVID-19 outcomes and vaccine uptake. For this study, we included participants who responded to at least one interview round starting October 2020 [V3].

The study was approved by the Institutional Review Board at the City University of New York (CUNY).

### Outcome definitions and ascertainment of vaccine hesitancy from October 2020-June 2021 and vaccine uptake as of July 2021

#### Vaccine hesitancy between October 2020 and June 2021

Using seven rounds of visits from October 2020 - June 2021, we assessed participants’ willingness to vaccinate with the question “If a coronavirus vaccine became available would you: a) Immediately get the vaccine; b) Delay getting the vaccine; c) Never get the vaccine.” Those who responded that they would “Delay getting vaccine” were categorized as *vaccine delayers* and those who responded that they would “Never get the vaccine” as *vaccine refusers*. COVID-19 vaccine delayers and refusers are together termed as COVID-19 vaccine hesitant, based on the WHO Strategic Advisory Group of Experts on Immunization (SAGE) definition of the term “vaccine hesitancy” (16). The outcome, “COVID-19 vaccine hesitancy,” was defined with three levels: vaccinate immediately, delay vaccine, and refuse vaccine. “Vaccinate immediately” was the reference category in statistical models. Participants who reported receiving the vaccine in a given interview were not asked about willingness to vaccinate again in subsequent interviews. We imputed their vaccine willingness status as “vaccinate immediately” for subsequent interviews assuming that if they received the vaccine, they were willing to get it immediately.

#### Vaccine uptake through July 2021

Since the vaccine became available to healthcare workers and high-risk individuals, in December 2020, we queried vaccination uptake (“Have you been vaccinated against COVID-19 with a Food and Drug Administration (FDA) approved vaccine/not in a vaccine trial: a) Yes; b) No; c) Don’t know/Not sure”) in all interview rounds starting December 2020. We also asked about vaccine-related side effects, motivation for getting vaccinated, and reasons for delay. Vaccine uptake through July 2021 was coded dichotomously (yes/no).

### Demographic characteristics

Participants’ age, race/ethnicity, income, education, and essential worker and healthcare worker status were determined at enrollment (V0, V1). Essential worker and healthcare worker status was asked again in October 2020 and the most recent reported status was used in the models. Self-reported race/ethnicity was coded based on standardized Office of Management and Budget categories (17).

### Ascertainment of COVID-19 related exposures and behavioral factors

To assess the association between prior exposure to COVID-19 and vaccine hesitancy, we defined COVID-19 history as a dichotomous variable using three inputs: self-reported COVID-19 PCR diagnosis or seropositivity (visits V0-V7), self-identifying as a COVID-19 long-hauler (visits V4-V7) or being seropositive for COVID-19 antibodies in two rounds of testing performed as part of our study from May-September 2020 and from November 2020-January 2021. To measure COVID-19 risk perception, we asked participants if they were worried that they would get sick from coronavirus, that their loved ones would get sick from coronavirus, and that coronavirus will overwhelm hospitals (not at all worried/ not too worried/ somewhat worried /very worried). COVID-19 related anxiety was measured using the Generalized Anxiety Disorder-7 (GAD-7) scale, and participants were categorized as having “no/low anxiety” or “moderate/high anxiety” based on their median scores (visits V0-V7). We also asked if they felt that the federal government was prioritizing the safety of citizens during the pandemic (agree/disagree/neutral) (V6).

To understand if vaccine hesitancy was correlated with the use of NPIs, we drew on participants’ responses to questions about their mask use and social distancing in public places in the June 2021 interview (V7) (Specific questions in Table 1 and sTable 2). We assigned a score of 1 for responses that indicated lack of participant engagement with individual NPIs and 0 otherwise. We summed the coded behaviors to create a risk score. Participants engaging in 5 or more risk-taking activities (median risk score=5) were defined as engaged in higher risk behavior. We separately assessed whether most recent masking behavior and air travel were associated with vaccine hesitancy in June 2021.

**Table 1:**
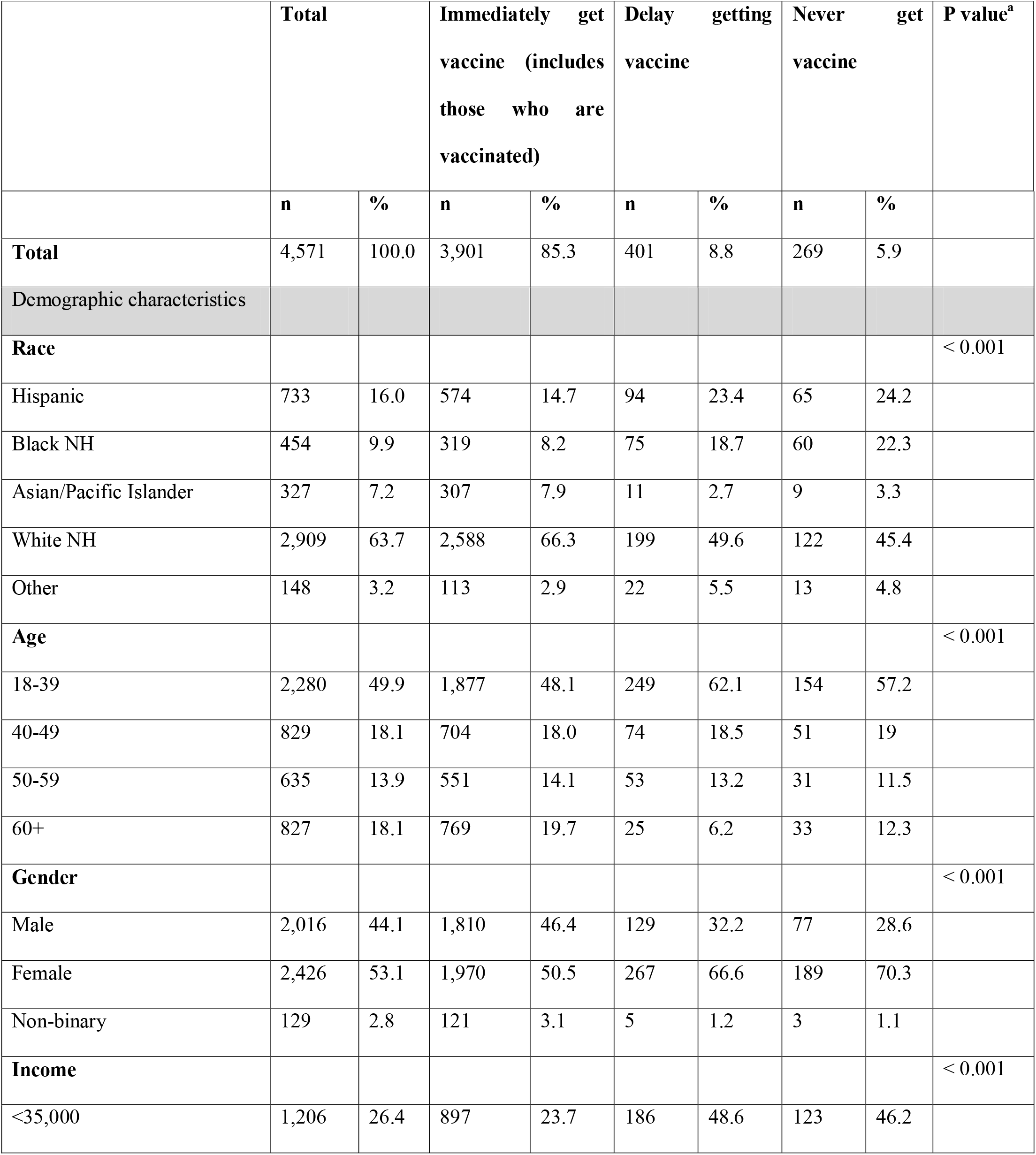
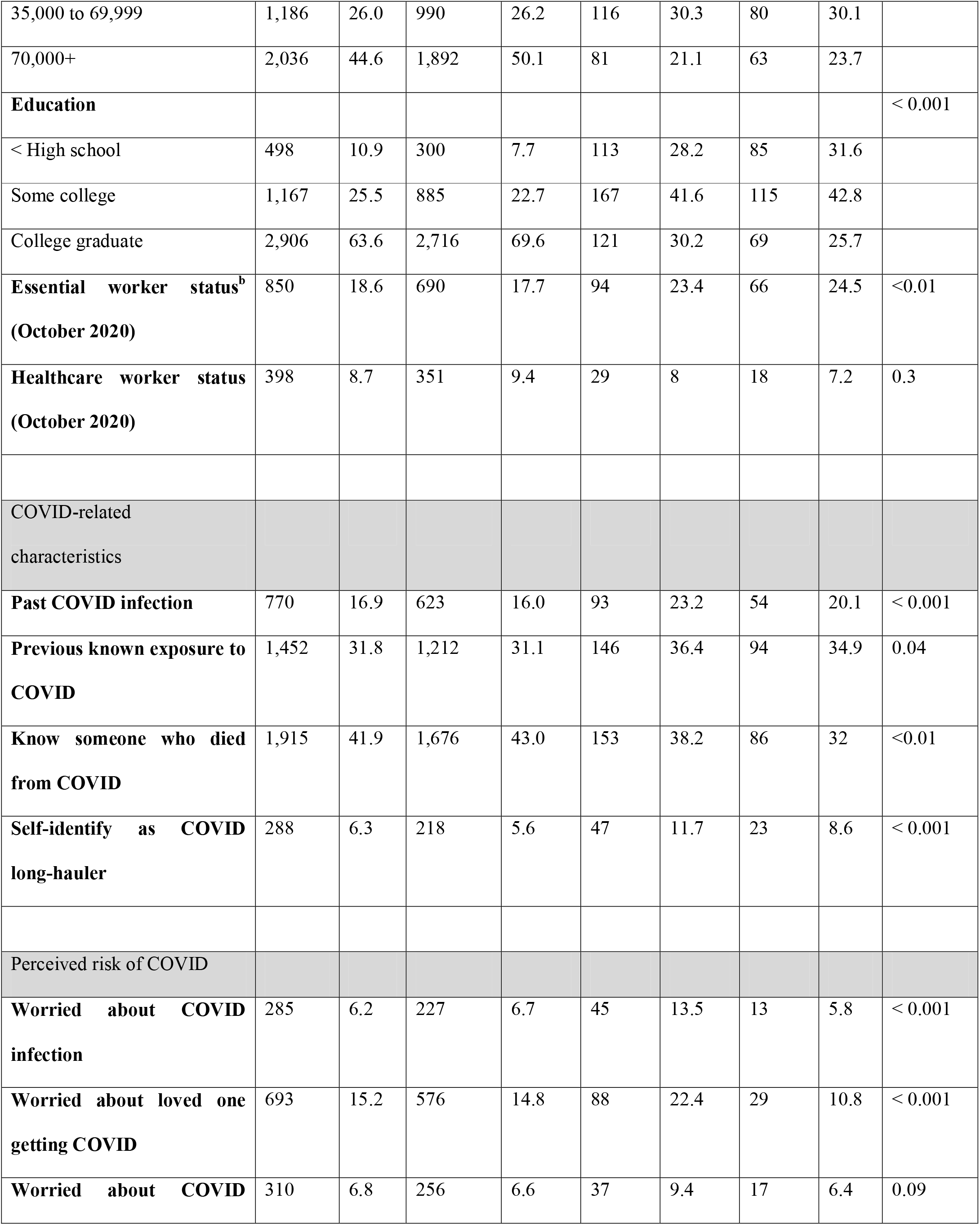
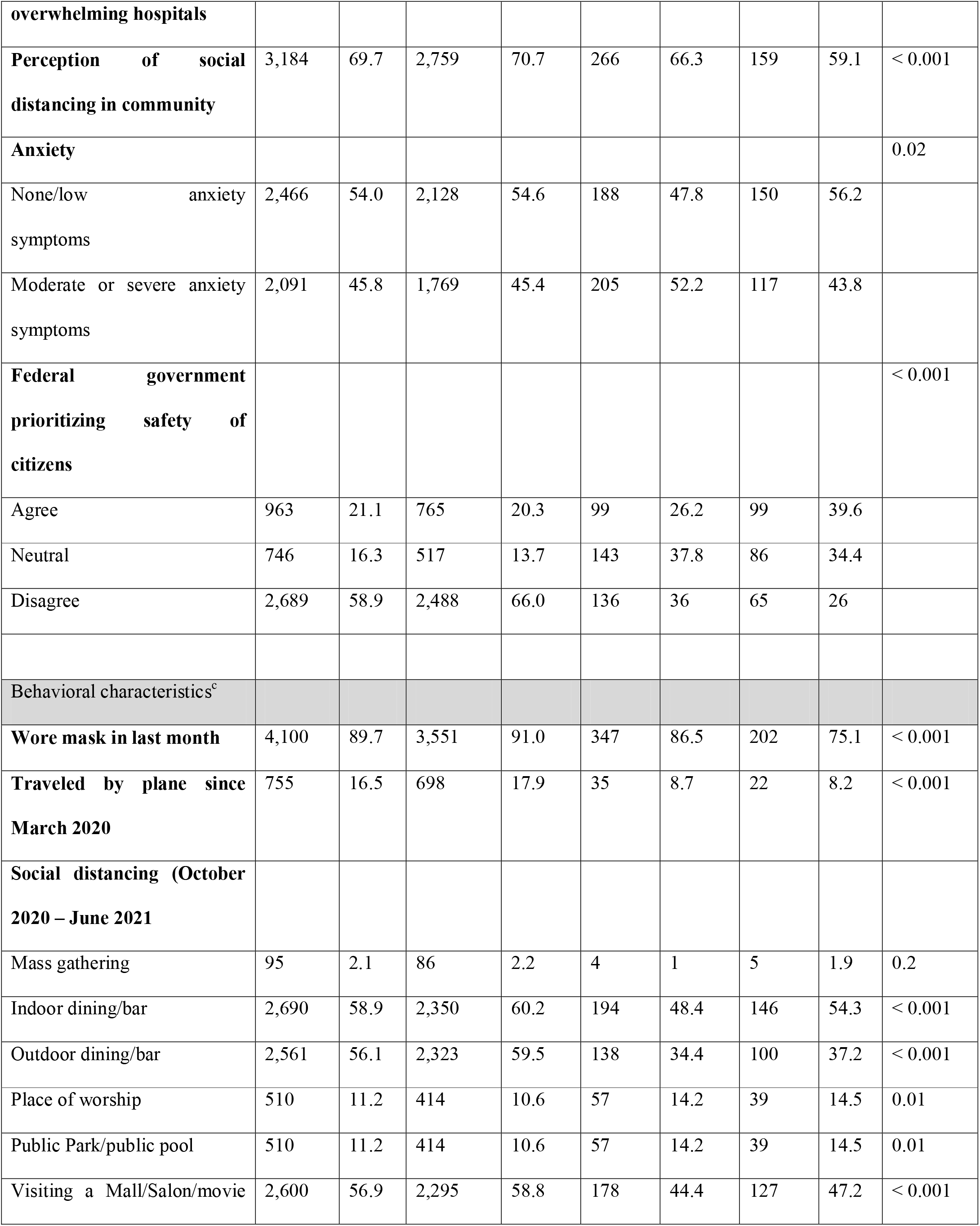
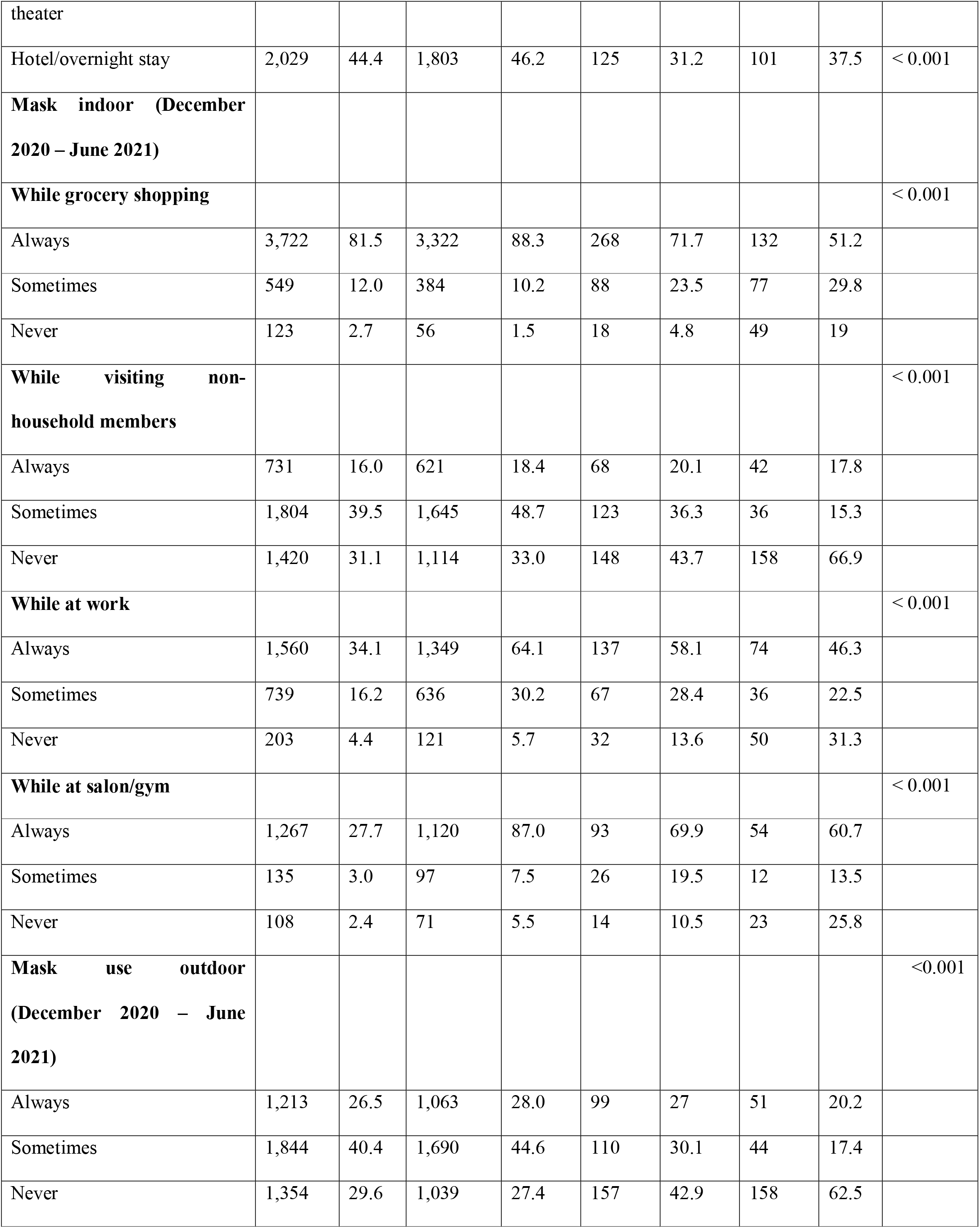
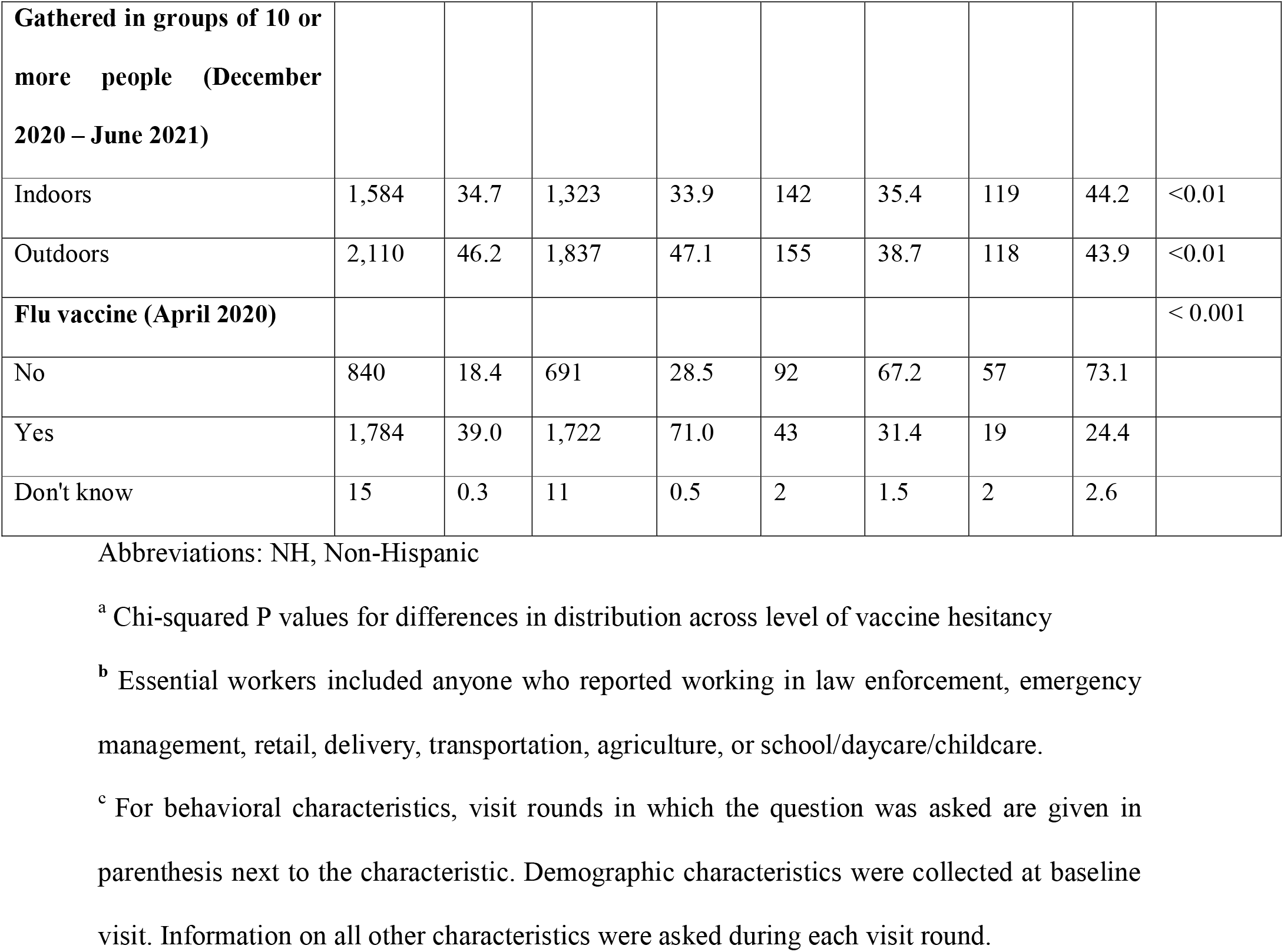
Cohort characteristics by vaccine hesitancy among CHASING COVID cohort participants in June 2021 (N=4,571)

### Statistical analysis

Chi-squared tests and corresponding *P* values were used to describe the distribution of patient characteristics across vaccine hesitancy levels in June 2021. Mean change in willingness to vaccinate over time was assessed using the McNemar-Bowker test. No imputation was performed for missing data and interview non-response was not associated with vaccination status. Models were implemented within the ‘multgee’ and ‘nnet’ packages in R version 4.0.1 (18).

#### Changes in willingness to vaccinate between October 2020 and July 2021

Multinomial generalized estimating equation (GEE) models were used to measure changes in vaccine hesitancy over time between October 2020 and July 2021 by estimating odds ratios (aORs) and 95% confidence intervals (CIs), adjusted for age, gender, race/ethnicity, and comorbidities. We included an interaction term between race/ethnicity and calendar time of interview in the model to test the hypothesis that the rate of change of vaccine hesitancy differed by race/ethnicity as found in some studies (19). Longitudinal correlation between participants was specified using an independence correlation matrix and variance was estimated using robust variance estimators.

#### Correlates of vaccine hesitancy in June 2021

Factors associated with vaccine hesitancy were assessed from the June 2021 interview (V7) using multinomial logistic regression to estimate ORs and 95% CIs. Separate models were built to assess the association between sociodemographic factors, COVID-19 history, behavioral characteristics, and COVID-19 risk perception and vaccine hesitancy. Models for COVID-19 history, behavioral characteristics, and COVID-19 risk perception were adjusted for sociodemographic factors that were statistically significantly associated with the outcome (2-sided *P* <0.05).

#### Association between vaccine hesitancy and subsequent vaccine uptake as of July 2021

Association between vaccine hesitancy and subsequent vaccine uptake was assessed using logistic regression models, which estimated ORs and 95% CIs. For those who received the vaccine, association with vaccine hesitancy was assessed using the most recent visit prior to the one when vaccine receipt was reported. Similarly, for those who remained unvaccinated as of July 2021, we used vaccine hesitancy status reported in the June 2021 visit (or prior if missing), to ensure exposure measurement in both groups is comparable. The model was adjusted for sociodemographic characteristics, comorbidities, and COVID-19 history.

## RESULTS

A total of 5,458 participants responded to at least one of the interview rounds between October 2020 and July 2021, of which 4,191 (76%) completed all full interview rounds. Vaccination status was obtained for all study participants and only one participant was missing vaccine willingness status across all visits. Vaccine delay decreased significantly from 51% to 6.7% between October 2020 to July 2021 (sTable 1). Decrease in vaccine refusal was also statistically significant but less prominent (8.4% to 5.7%). The Sankey plot (Figure 1) illustrates changes in vaccine hesitancy among participants. Most participants moved from the “delay” vaccine category to the “immediately” vaccinate category and eventually to the “vaccinated” category. Movement out of the “never” vaccinate category to “delay” or “immediately” vaccinate categories was limited.

**Figure 1:**
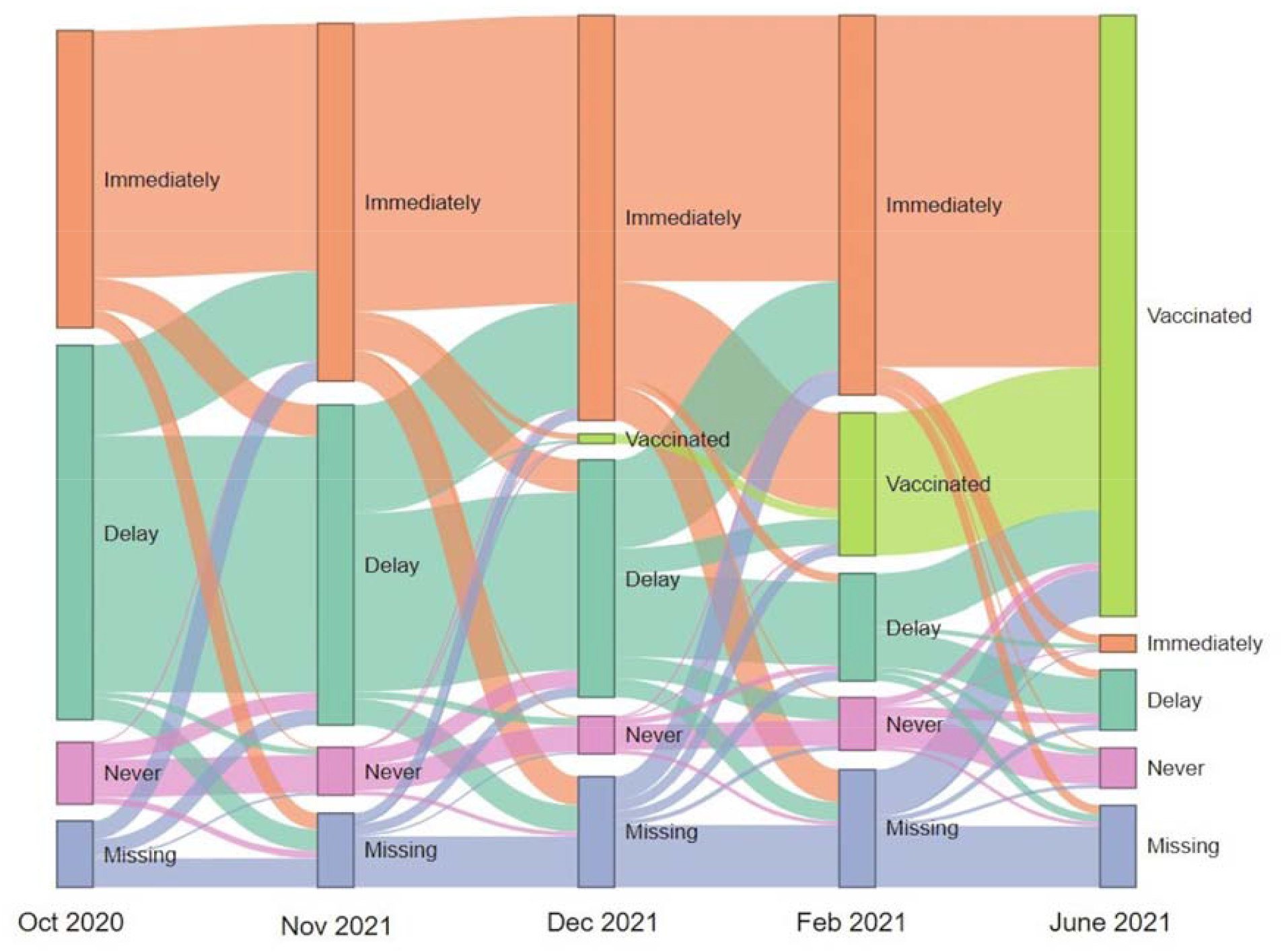
Sankey plot showing trends in vaccine hesitancy over time among CHASING COVID Cohort participants from October 2020 to June 2021. Number of participants who responded that they would immediately get the vaccine (orange), delay getting the vaccine (sage green), never get the vaccine (pink), or are already vaccinated (fluorescent green) when it becomes available to them are depicted in each rectangular node with the width of flows proportional to how many individuals report that response. Plot shows how their responses varied from October 2020 to June 2021. Information corresponding to only the full-format interviews is displayed here. Results from the short interviews conducted in April, May, and July 2021 not depicted here. Participants who responded to at least one of the interviews between October 2020 and June 2021 are included in this plot. Participants missing in a given interview round are shown in purple.

Of the cohort participants who responded to the most recent full interview (June 2021, n=4,571), 85.3% said they would immediately get the vaccine/were already vaccinated, 8.8% were vaccine delayers, and 5.9% were vaccine refusers (Table 1). Participants who were Hispanic and NH Black compared to NH White, younger (18-39 years old) compared to 60 years or older, female compared to male, had income < $35,000 compared to income >$70,000, and had less than high school education compared a college education were more likely to delay or refuse the COVID-19 vaccine.

### Trends in willingness to vaccinate between October 2020 and July 2021

Overall, vaccine delay decreased by 96% (aOR: 0.04, 95% CI: 0.03, 0.05) and vaccine refusal by 71% (aOR: 0.29, 95% CI: 0.24, 0.35) between October 2020 and July 2021, adjusted for race/ethnicity, age, gender, and comorbidities (Table 2). Even though vaccine hesitancy decreased overall, the rate of change differed by race/ethnicity. In December relative to October 2020, NH Black participants had 2.8 times (95% CI: 2.1, 3.6) higher odds of delaying vaccine compared to NH Whites, adjusted for age, gender and comorbidities (Figure 2). In July 2021 relative to October 2020, NH Black participants still had significantly higher odds of delaying COVID-19 vaccine than NH Whites (aOR:1.71; 95% CI: 1.15, 2.54). Similar trends in vaccine delay were seen for Hispanic compared to NH White participants. In December compared to October 2020, adjusted odds of vaccine refusal in NH Black participants were 1.6 times greater (95% CI: 1.15, 2.24) compared to NH Whites. However, by July 2021 compared to October 2020, vaccine refusal in NH Black participants was significantly lower than in NH Whites (aOR: 0.55, 95% CI: 0.55, 0.79), adjusted for age, gender and comorbidities. (Figure 2).

**Table 2:**
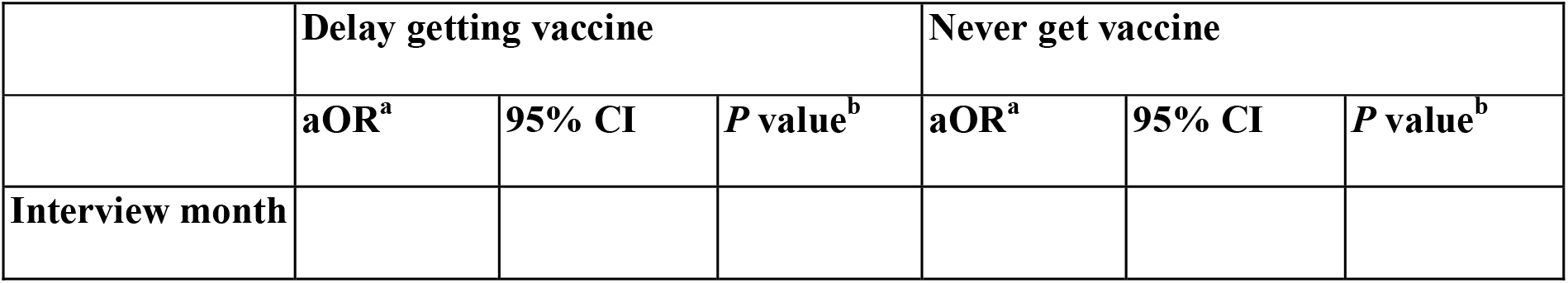
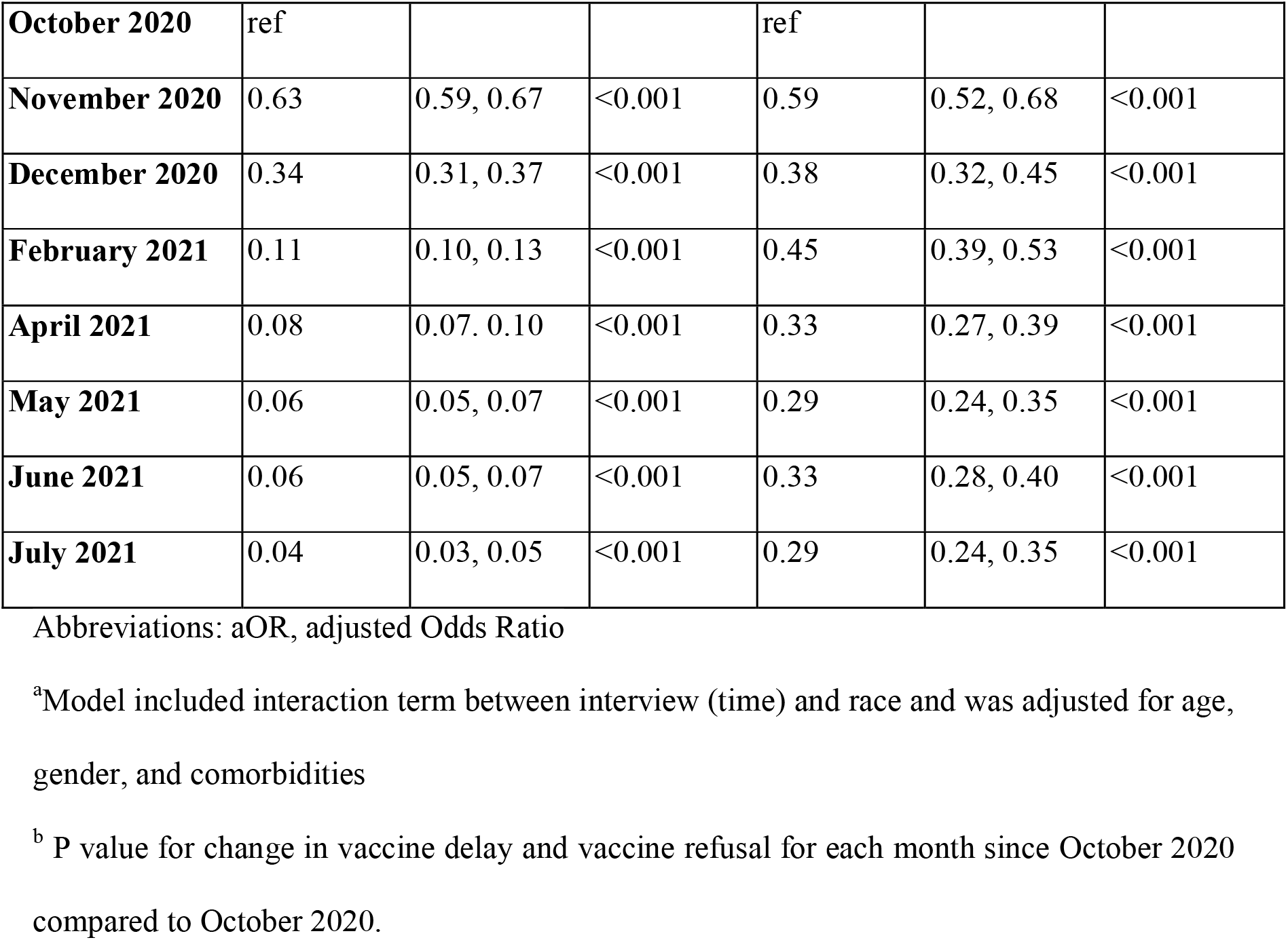
Generalized Estimating Equations (GEE) models for change in vaccine hesitancy over time among CHASING COVID cohort participants, October 2020-July 2021 (N=5,458)

|  | Delay getting vaccine |  |  | Never get vaccine |  |  |
| --- | --- | --- | --- | --- | --- | --- |
|  | aOR<sup>a</sup> | 95% CI | P value<sup>b</sup> | aOR<sup>a</sup> | 95% CI | P value<sup>b</sup> |
| **Interview month** |  |  |  |  |  |  |
| **October 2020** | ref |  |  | ref |  |  |
| **November 2020** | 0.63 | 0.59, 0.67 | <0.001 | 0.59 | 0.52, 0.68 | <0.001 |
| **December 2020** | 0.34 | 0.31, 0.37 | <0.001 | 0.38 | 0.32, 0.45 | <0.001 |
| **February 2021** | 0.11 | 0.10, 0.13 | <0.001 | 0.45 | 0.39, 0.53 | <0.001 |
| **April 2021** | 0.08 | 0.07, 0.10 | <0.001 | 0.33 | 0.27, 0.39 | <0.001 |
| **May 2021** | 0.06 | 0.05, 0.07 | <0.001 | 0.29 | 0.24, 0.35 | <0.001 |
| **June 2021** | 0.06 | 0.05, 0.07 | <0.001 | 0.33 | 0.28, 0.40 | <0.001 |
| **July 2021** | 0.04 | 0.03, 0.05 | <0.001 | 0.29 | 0.24, 0.35 | <0.001 |

Abbreviations: aOR, adjusted Odds Ratio
<sup>a</sup>Model included interaction term between interview (time) and race and was adjusted for age, gender, and comorbidities
<sup>b</sup> P value for change in vaccine delay and vaccine refusal for each month since October 2020 compared to October 2020.

**Figure 2:**
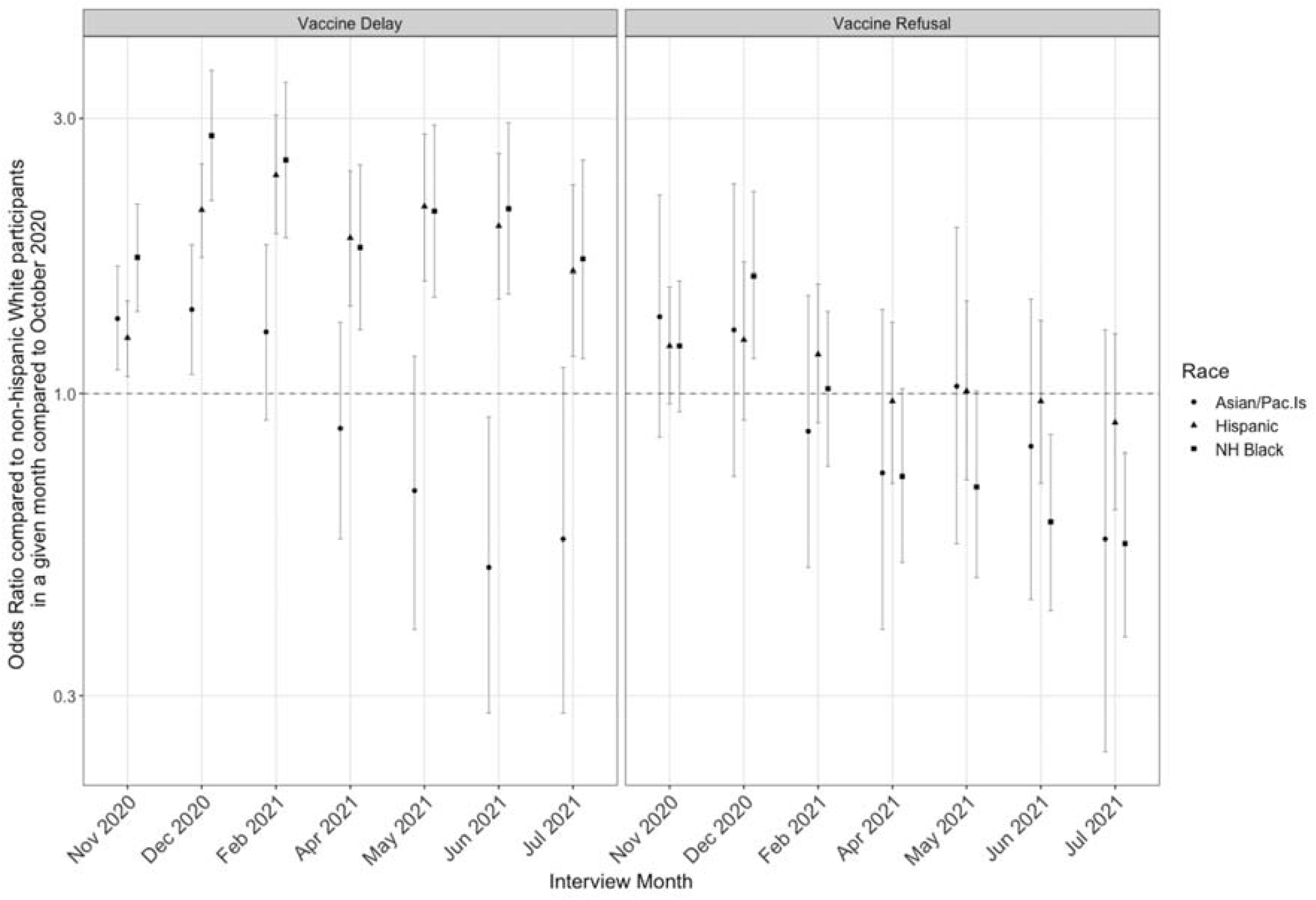
Racial/ethnic differences in change in vaccine hesitancy over time among CHASING COVID COHORT participants between October 2020 and July 2021. This plot shows odds ratios (ORs) of vaccine hesitancy for Asian/Pacific Islanders (circle), Hispanic (triangle), and NH Black (square) participants compared to NH White participants in each interview month compared to October 2020 (first interview when vaccine-related questions were asked). ORs for vaccine delay are in the left panel and vaccine refusal are in the right panel. Hispanic and NH Black participants had higher odds of vaccine delay compared to NH White in each subsequent visit after October 2020, with ORs being higher in the early vaccine era compared to later. However, odds of vaccine refusal for NH Black participants were lower compared to NH White participants in the recent visits compared to October 2020. This suggests a steeper decline in vaccine refusal among NH black participants compared to NH White participants since before vaccine roll-out. Pac.Is.: Pacific Islander; NH: non-Hispanic

### Correlates of vaccine hesitancy in June 2021

#### Sociodemographics

Compared to NH White participants, adjusted odds ratios for vaccine delay were 1.99 (95% CI: 1.47, 2.71) for NH Black, 1.29 (95% CI: 0.97, 1.71) for Hispanic, and 0.35 (95% CI: 0.18, 0.69) for Asian/Pacific Islander participants (Table 3, Model 1). Male and non-binary gender, older age, higher income, and college education were associated with lower odds of vaccine delay in the adjusted model.

**Table 3:** Multinomial logistic regression models for association between baseline characteristics and vaccine hesitancy among CHASIN COVID participants in June, 2021 (N= 4,571)

|  | Delay getting vaccine |  |  | Never get vaccine |  |  |
| --- | --- | --- | --- | --- | --- | --- |
|  | aOR | 95% CI | P value | aOR | 95% CI | P value |
| **Model 1: Baseline characteristics associated with vaccine hesitancy in December, 2020** |  |  |  |  |  |  |
| **Race/ethnicity** |  |  |  |  |  |  |
| NH White | ref |  |  | ref |  |  |
| NH Black | 1.99 | 1.47, 2.71 | <0.001 | 2.53 | 1.78, 3.59 | <0.001 |
| Hispanic | 1.29 | 0.97, 1.71 | 0.07 | 1.41 | 1.00, 1.96 | 0.04 |
| Asian/<br>Pacific<br>Islander | 0.35 | 0.18, 0.69 | 0.002 | 0.62 | 0.31, 1.22 | 0.16 |
| Other | 2.05 | 1.23, 3.43 | 0.005 | 1.88 | 0.99, 3.55 | 0.05 |
| **Gender** |  |  |  |  |  |  |
| Female | ref |  |  | ref |  |  |
| Male | 0.5 | 0.40, 0.63 | <0.001 | 0.48 | 0.36, 0.63 | <0.001 |
| Non-binary | 0.14 | 0.05, 0.39 | <0.001 | 0.17 | 0.05, 0.57 | 0.003 |
| **Age** |  |  |  |  |  |  |
| 18-39 | ref |  |  | ref |  |  |
| 40-49 | 0.86 | 0.65, 1.16 | 0.34 | 1.06 | 0.75, 1.49 | 0.71 |
| 50-59 | 0.85 | 0.61, 1.19 | 0.37 | 0.88 | 0.57, 1.34 | 0.55 |
| 60+ | 0.24 | 0.15, 0.38 | <0.001 | 0.65 | 0.43 0.99 | 0.04 |
| **Income** |  |  |  |  |  |  |
| <35,000 | ref |  |  | ref |  |  |
| 35,000 to 69,999 | 0.71 | 0.54, 0.92 | 0.009 | 0.78 | 0.56, 1.05 | 0.1 |
| 70,000+ | 0.42 | 0.31, 0.56 | <0.001 | 0.54 | 0.38, 0.76 | <0.001 |
| **Education** |  |  |  |  |  |  |
| < High school | ref |  |  | ref |  |  |
| Some college | 0.65 | 0.49, 0.87 | 0.004 | 0.54 | 0.39, 0.75 | <0.001 |
| College graduate | 0.21 | 0.15, 0.28 | <0.001 | 0.14 | 0.09, 0.19 | <0.001 |
| **Comorbidities<sup>a</sup>** |  |  |  |  |  |  |
| No | ref |  |  | ref |  |  |
| Yes | 0.74 | 0.58, 0.94 | 0.01 | 0.62 | 0.47, 0.83 | 0.001 |
| **Model 2<sup>b</sup>: Essential worker status and vaccine hesitancy** |  |  |  |  |  |  |
| **Essential workers<sup>c</sup>** | 1.11 | 0.85, 1.46 | 0.41 | 1.17 | 0.86, 1.60 | 0.3 |
| **Model 3<sup>b</sup>: HCW status and vaccine hesitancy** |  |  |  |  |  |  |
| **Healthcare workers** | 0.7 | 0.46, 1.06 | 0.09 | 0.65 | 0.39, 1.09 | 0.1 |

Abbreviations: aOR, adjusted Odds Ratio; NH, Non-Hispanic
<sup>a</sup> Comorbidity is defined as having history of heart attack, depression, angina, immunosuppression, type 2 diabetes, high blood pressure, cancer, asthma, COPD, chronic kidney disease, and/or HIV/AIDS
<sup>b</sup> Models 2 and 3 adjusted for baseline characteristics and past COVID-19 history
<sup>c</sup> Essential workers included anyone who reported working in law enforcement, emergency management, retail, delivery, transportation, agriculture, or school/daycare/childcare.

### COVID-19 experience

Those who knew someone who died from COVID-19 had significantly lower odds of refusing COVID-19 vaccine (aOR: 0.60, 95% CI: 0.46, 0.80) compared to those who did not, after adjusting for demographic factors. Participants who self-identified as COVID long-haulers had significantly higher odds of being vaccine delayers (aOR: 1.62, 95% CI: 1.14, 1.29) (sTable 2). Prior COVID-19 diagnosis and serostatus were not associated with vaccine hesitancy.

Worrying about themselves or their loved ones getting COVID-19 was associated with lower odds of vaccine refusal (aOR: 0.48, 95% CI: 0.33, 0.71). Those who reported suffering from moderate to severe COVID-related anxiety had lower odds of delaying vaccine (aOR: 0.79, 95% CI: 0.63, 1.00) and vaccine refusal (aOR: 0.62, 95% CI: 0.47, 0.82). Those who did not trust the federal government to prioritize the safety of citizens during the pandemic (as measured in the May 2021 survey) had lower odds of delaying the vaccine (aOR: 0.53, 95% CI: 0.40, 0.71) or refusing it (aOR: 0.25, 95% CI: 0.18, 0.35) (sTable 2).

### Behaviors

Participants who reported wearing a mask in the prior month had substantially lower odds of vaccine refusal (aOR: 0.42, 95% CI: 0.31, 0.58) (sTable 2). Air travel was also associated with a lower odds of vaccine delay (aOR: 0.50, 95% CI: 0.34, 0.72) and vaccine refusal (aOR:0.50, 95% CI: 0.32, 0.79). Those who engaged in a greater number of high-risk activities (such as not wearing masks consistently in public areas, not maintaining social distancing with non-household members, gathering in large groups) had higher odds of refusing the COVID-19 vaccine (aOR: 1.79, 95% CI: 1.37, 2.34).

### Association between vaccine hesitancy and subsequent vaccine uptake through July 2021

A total of 4,197 (76.9%) participants had reported receiving at least one dose of the coronavirus vaccine as of July 2021 (visit V7.1). Those who reported they would delay vaccination in the previous interview had 85% lower odds of receiving the vaccine immediately (aOR: 0.15, 95% CI: 0.13, 0.18) while those who reported they would never get vaccinated had 98% lower odds (aOR: 0.02; 95% CI: 0.01, 0.03) of being vaccinated as of July 2021, adjusted for demographic factors, COVID-19 history, and comorbidities. Compared to NH White participants, NH Black participants had lower odds of being vaccinated (aOR: 0.71, 95% CI: 0.56, 0.91). Among those who were willing to vaccinate, 7% of NH White and 6% of Asian/Pacific Islander participants had not yet received a vaccine in July 2021, compared to 19% of NH Black participants and 13% of Hispanic participants. Older age, higher income, higher education, residence in the Midwest or the Northeast of the U.S. compared to the South, having comorbidities, and having a history of COVID-19 were associated with higher odds of being vaccinated (Table 4).

**Table 4:** Association between vaccine hesitancy and vaccine uptake among CHASING COVID participants as of July, 2021(N=5,458)

|  | COVID-19 Vaccine uptake |  |  |
| --- | --- | --- | --- |
|  | aOR<sup>a</sup> | 95% CI | P value |
| **Covid-19 Vaccine hesitancy reported in the visit prior to receiving vaccine** |  |  |  |
| Immediately | ref |  |  |
| Delay | 0.02 | 0.01, 0.03 | <0.001 |
| Never | 0.15 | 0.13, 0.18 | <0.001 |
| **Race/ethnicity** |  |  |  |
| NH White | ref |  |  |
| Hispanic | 0.94 | 0.76, 1.17 | 0.62 |
| NH Black | 0.71 | 0.56, 0.91 | 0.007 |
| Asian | 1.39 | 0.98, 1.99 | 0.06 |
| Other | 0.58 | 0.39, 0.88 | 0.009 |
| **Age** |  |  |  |
| 18-39 | ref |  |  |
| 40-49 | 1.12 | 0.90, 1.39 | 0.27 |
| 50-59 | 1.33 | 1.02, 1.74 | 0.03 |
| 60+ | 1.64 | 1.25, 2.14 | <0.001 |
| **Gender** |  |  |  |
| Female | ref |  |  |
| Male | 0.96 | 0.81, 0.13 | 0.68 |
| Non-binary | 1.31 | 0.78, 0.30 | 0.31 |
| **Income** |  |  |  |
| <35,000 | ref |  |  |
| 35,000 to 69,999 | 1.34 | 1.09, 1.65 | 0.004 |
| 70,000+ | 1.8 | 1.45, 2.23 | <0.001 |
| **Education** |  |  |  |
| < High school | ref |  |  |
| Some college | 1.28 | 1.01, 1.64 | 0.04 |
| College graduate | 2.34 | 1.82, 2.99 | <0.001 |
| **Region** |  |  |  |
| South | ref |  |  |
| Midwest | 1.44 | 1.13, 1.82 | 0.002 |
| Northeast | 1.38 | 1.12, 1.71 | 0.002 |
| West | 1.25 | 1.01, 1.54 | 0.04 |
| **Any comorbidity<sup>b</sup>** |  |  |  |
| No | ref |  |  |
| Yes | 1.33 | 1.11, 1.58 | 0.001 |
| **COVID-19 history<sup>c</sup>** |  |  |  |
| No | ref |  |  |
| Yes | 1.29 | 1.05, 1.58 | 0.01 |
| **Children under 18 years of age in household** |  |  |  |
| No | ref |  |  |
| Yes | 0.56 | 0.46, 0.68 | <0.001 |

Abbreviations: aOR, adjusted Odds Ratio; NH, Non-Hispanic
<sup>a</sup>Odds ratios for vaccine uptake were adjusted for age, race/ethnicity, gender, income, education, region of residence, past COVID-19 history, presence of children <18 in the household, and comorbidities
<sup>b</sup> Comorbidity was defined as having history of heart attack, depression, angina, immunosuppression, type 2 diabetes, high blood pressure, cancer, asthma, COPD, chronic kidney disease, and/or HIV/AIDS
<sup>c</sup> COVID-19 history is defined as someone who had a PCR diagnosis, self-identified as a long hauler, or was seropositive

Among unvaccinated participants who reported they would delay or never get the vaccine, the most frequently cited reasons for vaccine delay in June 2021 were concerns about long-term side effects (26.7%), short-term side effects (18.9%), and concerns about vaccine effectiveness (15.9%). Reasons for delay did not vary by race/ethnicity. Among those willing to take the vaccine immediately, the majority (17.9%) responded that they wanted to be vaccinated to avoid getting COVID-19, end the pandemic (14.4%), protect themselves (15.6%), and protect others (13.8%).

## DISCUSSION

After a rapid roll out in spring 2021, the pace of COVID-19 vaccination uptake in the U.S. has slowed down (6). Using a prospective cohort study, we tracked how COVID-19 vaccine hesitancy evolved as vaccine became widely available, factors associated with vaccine hesitancy, and the association between intention to vaccinate and subsequent vaccine uptake. We found that while the proportion of vaccine delayers decreased significantly in our cohort as vaccine availability improved, vaccine refusal remained relatively constant. Vaccine hesitancy differed by race/ethnicity, age, income, and education. While vaccine delay decreased overall, it did so at a slower rate for participants of color compared to NH White participants since October 2020. Conversely, there has been a greater decrease in vaccine refusal over time among NH Black compared to NH White participants. Willingness to vaccinate was strongly associated with subsequent vaccine uptake in our cohort.

We separately assessed correlates of COVID-19 vaccine delay and vaccine refusal. This delineation is important because the factors that drive vaccine delay and refusal could be different (20,21), especially for the COVID-19 vaccine, and public health messaging will need to be tailored to each group to address their specific concerns. Similar to other U.S. based surveys, we found age, sex, race/ethnicity, income, and education to be correlated with vaccine hesitancy (11,22–24). Even after large scale roll-out, as of July 2021 vaccine uptake was lower among NH Black participants, 18–39-year-olds, those with income lower in < $35,000, and those with only a high school education. Communities of color, low-income groups, and those with fewer years of education have experienced particularly high COVID-19 infection rates, hospitalization rates and mortality rates (25–28). Given that vaccine uptake remains low in these groups, they may remain more susceptible to a higher COVID-19 burden, especially as more transmissible variants such as B.1.617.2 (delta) continue to emerge (29,30).

Several studies have focused on racial/ethnic differences in COVID-19 vaccine hesitancy, and our findings are consistent with these studies (11,31–34). Even though overall vaccine delay decreased with time, racial/ethnic gaps in vaccine delay persisted from November 2020 to July 2021. Distrust in the medical community may be an important reason. Historical mistreatment, oppression, and unethical conduct from the government, medical establishments, and scientific research communities have adversely impacted racial and ethnic minorities, especially Black Americans (31,35,36). Recognizing this, healthcare professionals have sought to boost vaccine confidence to implement a more equitable vaccination campaign in the U.S. (37). Interestingly, we saw that outright vaccine refusal among NH Black participants declined to a much greater degree compared to NH White participants as vaccines became widely available, suggesting that these targeted efforts in minority communities might have been successful in changing the minds of even those who said they would deny the vaccine at a time when vaccines were under development.

Similar to other studies, we found racial disparities in vaccine uptake (34,38). Even among participants willing to vaccinate, a higher proportion of NH White and Asian participants reported being vaccinated compared to Hispanic and NH Black participants, which may indicate barriers to vaccine access. One study in New York City found fewer vaccination sites in districts with higher poverty rates and higher proportions of Black and Latinx residents (39). Racial/ethnic minorities constitute a large proportion of the essential workforce (40) which might make it difficult for them to take time off work for vaccination and or recover from potential vaccine side-effects. It is critical that testing and vaccinations are accessible and without cost barriers to reduce the disproportionately high COVID-19 burden in communities of color. Employers can encourage vaccination among workers by offering on-site vaccine drives, providing paid sick leave, and paid transportation to and from vaccine sites. Reduced waiting times and easy availability of the vaccine in health centers, pharmacies, or at home might further improve vaccination rates (41).

In our cohort, COVID-19 long-haulers reported being more likely to delay vaccination, perhaps due to fears that vaccination might exacerbate their symptoms. While the CDC has no specific guidelines regarding vaccinations for COVID-19 long-haulers, it recommends that people get vaccinated regardless of their infection history because infection-derived immunity may wane over time (42,43). Early evidence suggests COVID-19 vaccines have the potential to resolve long-COVID symptoms and large clinical studies are being undertaken to study this (44–46). Cohort participants who were less likely to engage in protective behavior such as mask-wearing and social distancing were also less willing to be vaccinated. This may be due to lack of trust in authorities as source of COVID-19 information (47). Unvaccinated individuals who do not engage in NPIs could be at an even higher risk of COVID-19 infection and mortality.

The main stated reasons for COVID-19 vaccine hesitancy in our cohort was worry about short-term and long-term vaccine side-effects, and concerns about vaccine effectiveness. Studies show that targeted informational interventions may not overcome vaccine hesitancy (48,49), suggesting more research is needed to effectively communicate vaccination benefits and address concerns about risks to those who remain hesitant. Being transparent about vaccine risks and uncertainties around new vaccines and emerging virus strains may help increase public trust in government bodies (50,51). Encouragingly, the proportion of vaccine delayers decreased over time, perhaps due to new information about vaccine safety and efficacy emerged and an increasing number of people got vaccinated without incident. To assuage concerns about effectiveness and safety, studies that estimate direct and population-level vaccine effects for the different COVID-19 vaccines and in different subpopulations using real-world data in the U.S. are critical (52–54). Powering randomized control trials to increase the chances of detecting less common side effects may also help.

The strengths of our study include prospective assessment of association between willingness to vaccinate and subsequent vaccine uptake, a diverse and geographically representative cohort, and detailed data on demographics, biomarkers on prior COVID-19 exposures, and behavioral characteristics that are not available in surveillance databases, electronic medical records, or cross-sectional surveys/polls. Our prospective design with repeat measurements allowed us to assess how individual-level vaccine hesitancy has changed over the course of the pandemic as vaccines became more widely available.

Our study also has limitations. Participants self-reported vaccination status as well as exposures, so the study is subject to misclassification and reporting bias. Because enrollment was done online, those without smartphones, computers, or a stable internet connection were less likely to be included. This is not a population-based sample as participants had to opt into the study. While we adjusted all models for demographic factors, there is a possibility of unmeasured and/or uncontrolled confounding. Not all participants responded to all interview rounds, although visit-level missingness was not associated with vaccination status. However, missingness was associated with age, sex, college education, serostatus, and COVID-19 history, which could potentially bias our results.

In summary, the proportion of COVID-19 vaccine delayers decreased substantially from October 2020 to July 2021 as vaccine availability increased. However, a small fraction (5.7%) of participants continue to refuse the vaccine. While racial/ethnic disparities in vaccine delay and vaccine uptake persisted, decrease in vaccine refusal was greater for racial minorities compared to NH Whites over time. To address disparities in vaccine uptake, awareness efforts and response to vaccine-related concerns should focus on low-income individuals and racial/ethnic minority individuals, and vaccine availability should be prioritized in communities where uptake has been low. To mitigate the impact of COVID-19 as a public health threat, it is important that no groups are left behind by vaccination initiatives.

## Data Availability

All data referenced in the manuscript are available upon request.

## Acknowledgements

We thank Dr. Patrick Sullivan and MTL labs for local validation work on the serologic assays for use with dried blood spots that greatly benefited our study. We are also grateful to MTL Labs for processing specimen collection kits and serologic testing of our cohort’s specimens.

## Preprint

The authors will upload this version of the article on Medrxiv (link not yet available). An earlier preprint version of the article can be found here: https://www.medrxiv.org/content/10.1101/2021.05.12.21257116v1

## Abbreviations

COVID-19: coronavirus disease-19
WHO: World Health Organization
US: United States
mRNA: (messenger ribonucleic acid)
NH: Non-Hispanic
C3 study: Communities, Households, and SARS-CoV-2 Epidemiology (CHASING) COVID Cohort study
SAGE: Strategic Advisory Group of Experts on Immunization
FDA: Food and Drug Administration
GAD-7): Generalized Anxiety Disorder-7
NPI: Non-Pharmaceutical Interventions
GEE: generalized estimating equations
aOR: adjusted Odds Ratios
CI: Confidence Intervals

