## Supplemental Material for "Determinants and trends of COVID-19 vaccine hesitancy and vaccine uptake in a national cohort of U.S. adults: A longitudinal study"

sTable 1: Trends in vaccine hesitancy over time among CHASING COVID participants, Octobe 2020 – July 2021

|  | **Total** | **Immediately get vaccine** | | **Delay getting vaccine** | | **Never get vaccine** | | **% delayers** | **% refusers** | **P value^a^** |
| --- | --- | --- | --- | --- | --- | --- | --- | --- | --- | --- |
|  | **n** | **n** | **%** | **n** | **%** | **n** | **%** |  |  |  |
| **October 2020** | 5,045 | 2,044 | 40.5 | 2,575 | 51 | 426 | 8.4 | 51 | 8.4 |  |
| **November 2020** | 5,028 | 2,475 | 49.2 | 2,221 | 44.2 | 332 | 6.6 | 44.2 | 6.6 | < 0.0001 |
| **December 2020** | 4,565 | 2,726 | 59.7 | 1,595 | 34.9 | 244 | 5.3 | 34.9 | 5.3 | < 0.0001 |
| **February 2021** | 4,649 | 3,556 | 76.5 | 734 | 15.8 | 359 | 7.7 | 15.8 | 7.7 | < 0.0001 |
| **June 2021** | 4,571 | 3,901 | 85.3 | 401 | 8.8 | 269 | 5.9 | 8.8 | 5.9 | < 0.0001 |
| **July 2021** | 3,882 | 3,402 | 87.6 | 259 | 6.7 | 221 | 5.7 | 6.7 | 5.7 | < 0.0001 |

a McNemar-Bowker test P values comparing vaccine delay and refusal trends in in each month since October 2020 to July 2021

sTable 2: Multinomial logistic regression models for association between vaccine hesitancy and COVID-19 history, risk perception, and risk behavior among CHASING COVID cohort participants in June 2021

|  | **Delay getting vaccine** | |  | **Never get vaccine** | |  |
| --- | --- | --- | --- | --- | --- | --- |
|  | **aOR^a^** | **95% CI** | ***P* value** | **aOR^a^** | **95% CI** | ***P* value** |
| **COVID-19 history^b^** | 1.22 | 0.94, 1.59 | 0.12 | 0.98 | 0.70, 1.35 | 0.9 |
| **Prior PCR diagnosis** | 0.93 | 0.67, 1.30 | 0.7 | 1.04 | 0.71, 1.52 | 0.81 |
| **Know someone who died from COVID** | 0.81 | 0.65, 1.02 | 0.08 | 0.6 | 0.46, 0.80 | <0.001 |
| **Long-hauler** | 1.62 | 1.14, 1.29 | 0.005 | 1.09 | 0.69, 1.73 | 0.69 |
| **Serostatus** |  |  |  |  |  |  |
| Ever positive | ref |  |  | ref |  |  |
| Negative | 0.81 | 0.5, 1.32 | 0.4 | 1.02 | 0.54, 1.92 | 0.94 |
| No test | 1.18 | 0.71, 1.93 | 0.51 | 1.48 | 0.78, 2.80 | 0.22 |
| **Worried about COVID** | 0.98 | 0.75, 1.30 | 0.93 | 0.48 | 0.33, 0.71 | <0.001 |
| **Perception of SD in community** | 1 | 0.79, 1.26 | 0.97 | 0.74 | 0.56, 0.96 | 0.03 |
| **Anxiety** |  |  |  |  |  |  |
| None/Low anxiety | ref |  |  | ref |  |  |
| Moderate/severe anxiety | 0.79 | 0.63, 1.00 | 0.05 | 0.62 | 0.47, 0.82 | <0.001 |
| **Trust that federal government is prioritizing safety of citizens during the pandemic** |  |  |  |  |  |  |
| Agree | ref |  |  | ref |  |  |
| Neutral | 1.69 | 1.25, 2.29 | <0.001 | 0.97 | 0.69, 1.35 | 0.86 |
| Disagree | 0.53 | 0.40, 0.71 | <0.001 | 0.25 | 0.18, 0.35 | <0.001 |
| **Wore a mask in the last month** | 0.97 | 0.70, 1.35 | 0.8 | 0..42 | 0.31, 0.58 | <0.001 |
| **Air travel** | 0.5 | 0.34, 0.72 | <0.001 | 0.5 | 0.32, 0.79 | 0.003 |
| **High risk behavior**^c^ **(Score >4)** | 0.9 | 0.72, 1.14 | 0.4 | 1.79 | 1.37, 2.34 | <0.001 |

Abbreviations: aOR, adjusted Odds Ratio; PCR, Polymerase chain reaction

^a^Models adjusted for baseline characteristics

**^b^** COVID-19 history is defined as someone who had a PCR diagnosis, self-identified as a long-hauler, or was seropositive

^c^ High risk behavior: Participants engaging in 5 or more risk-taking activities are considered high risk. Risk taking activities include not social distancing in public places, not wearing masks in grocery stores, at work, in other peoples' households, in gyms, outdoors, gathering in groups of >10 outdoors or indoors.
